# Viraemia and HIV drug resistance among people receiving dolutegravir versus efavirenz-based first-line antiretroviral therapy

**DOI:** 10.1101/2023.06.16.23291523

**Authors:** Jienchi Dorward, Yukteshwar Sookrajh, Richard Lessells, Elliot Bulo, Nicola Bodley, Lavanya Singh, Pravikrishnen Moodley, Natasha Samsunder, Paul K. Drain, Gail Hayward, Christopher C Butler, Nigel Garrett

## Abstract

Limited data exists to inform management of viraemia among people receiving dolutegravir-based first-line ART in low- and middle-income countries. Among South-Africans with viraemia ≥1000copies/mL receiving dolutegravir (n=43) and efavirenz (n=37), we found no dolutegravir resistance, but high efavirenz resistance (66.7%). 12-week resuppression was higher with dolutegravir (85%) versus efavirenz (38%).

## INTRODUCTION

Dolutegravir, an integrase strand transfer inhibitor (INSTI), is currently being rolled out across low and middle income countries (LMICs).^1 2^ It has shown better effectiveness, tolerability, and has a higher genetic barrier to drug resistance compared to previous non-nucleoside reverse transcriptase (NNRTI)-based regimens such as efavirenz.^3^ People with viraemia receiving dolutegravir may be more likely to have inconsistent adherence, than HIV drug resistance. However, the absence of widespread HIV drug resistance testing in LMICs,^4^ makes it difficult for clinicians to determine the cause of viraemia and manage it appropriately.

Among people receiving NNRTIs with viral failure (two consecutive viral loads [VLs] ≥1000 copies/mL, ≥3 months apart), approximately 70% have drug resistance, and therefore current World Health Organization (WHO) guidelines recommend switching to second-line ART^5 6^. Current WHO guidelines for managing viraemia on first-line dolutegravir are less clear, because there is little data from LMICs regarding dolutegravir drug resistance and subsequent VL outcomes.

Therefore, among people with viraemia on dolutegravir and efavirenz-based first-line ART, we aimed to compare subsequent VL trajectories and drug resistance profiles.

## METHODS AND ANALYSIS

We used data from the POwER study, a randomised study of point-of-care VL testing among people with HIV viraemia receiving first-line ART. The protocol and results have been previously published^7 8^.

### Setting and participants

POwER was conducted at two public clinics in KwaZulu-Natal, South Africa, where dolutegravir has been recommended for first- and second-line ART from December 2019.^9^ People with viraemia ≥1000 copies/mL are recommended to receive enhanced adherence counselling, with a repeat three month VL. If this remains high, those receiving efavirenz are recommended to switch to second-line ART, while those receiving dolutegravir should continue enhanced adherence counselling and repeat VL testing. Eligibility criteria for POwER were being ≥18 years old, non-pregnant, and receiving first-line dolutegravir or efavirenz-based ART, with viraemia ≥1000 copies/mL in the past six weeks and yet to receive enhanced adherence counselling. Dolutegravir recipients may have been initiated on dolutegravir, or previously transitioned from efavirenz.

## Procedures

Consenting participants were enrolled, received enhanced adherence counselling,^10^ and were randomised to point-of-care or standard laboratory based VL testing after 12 weeks. Management of these VL results and clinical care during the 24 weeks of follow-up was provided by public sector healthcare workers. Plasma samples were stored at enrolment, 12 week VL and 24 week study exit visits, for retrospective VL and drug resistance testing, with results not used for clinical management. All samples with VL ≥500 copies/mL were sequenced using next-generation sequencing with the Illumina MiSeq platform (Illumina, San Diego, CA) (see supplementary material). We identified drug resistance mutations (DRM) at >20% frequency in protease, integrase and reverse transcriptase regions using the Stanford HIVDR database.

### Variables and analyses

The main exposure was dolutegravir- or efavirenz-based ART at enrolment. We conducted descriptive analyses and used Fisher’s exact test to assess the proportions in each ART group who had viraemia ≥1000 copies/mL at enrolment, 12 weeks and 24 weeks. We also assessed the proportions with HIV drug resistance at each timepoint, and switched to second-line ART.

### Ethical approvals

The University of KwaZulu-Natal Biomedical Research Ethics Committee (BREC 00000836/2019) and the University of Oxford Tropical Research Ethics Committee (OxTREC 66-19) approved the study.

## RESULTS

### Participants

We enrolled 80 eligible participants between August-March 2022. Median age was 38.5 years (interquartile range [IQR] 33-45), 58.8% were female, and median time on ART was 3.2 years (IQR 1.0-6.0) (Table S1).

At enrolment, 37 (46.3%) had been receiving efavirenz-based first-line regimens for a median of 3.2 years (1.1-5.0), and 43 (53.7%) had been receiving dolutegravir for a median of 0.7 years (IQR 0.5-1.1). 15/43 (34.9%) had been initiated on dolutegravir, while the other 28/43 (65.1%) had been initiated on an efavirenz-based regimen and were subsequently transitioned to first-line dolutegravir. The dolutegravir group had less time on ART, slightly higher incomes and higher CD4 counts, but otherwise were similar to the efavirenz group (Table S1). All participants were receiving tenofovir disoproxil fumarate, apart from one dolutegravir participant receiving abacavir.

### Viraemia and HIV drug resistance

#### Enrolment

The median time since the pre-enrolment VL of ≥1000 copies/mL to enrolment was around two weeks (Table 1). At enrolment, the numbers with viraemia ≥1000 copies/mL had fallen to 18/43 (41.9%) dolutegravir participants, compared to 27/37 (73.0%) efavirenz participants (p=0.007). Of the 50 participants with VLs >500 copies/mL, HIVDR testing was successful in 48 for reverse transcriptase and 47 for integrase. The proportion with DRMs against either of the nucleoside reverse transcriptase inhibitor (NRTI) backbone drugs was lower in dolutegravir participants (2/19, 10.5%, 95% CI 1.9, 32.9) compared to efavirenz participants (21/29, 72.4%, 54.0, 85.4, p=<0.001, Table 1). In efavirenz participants, 25/29 (86.2%, 68.7, 95.0) had DRMs against efavirenz, while among dolutegravir participants there were no DRMs against dolutegravir.

**Table 1:**
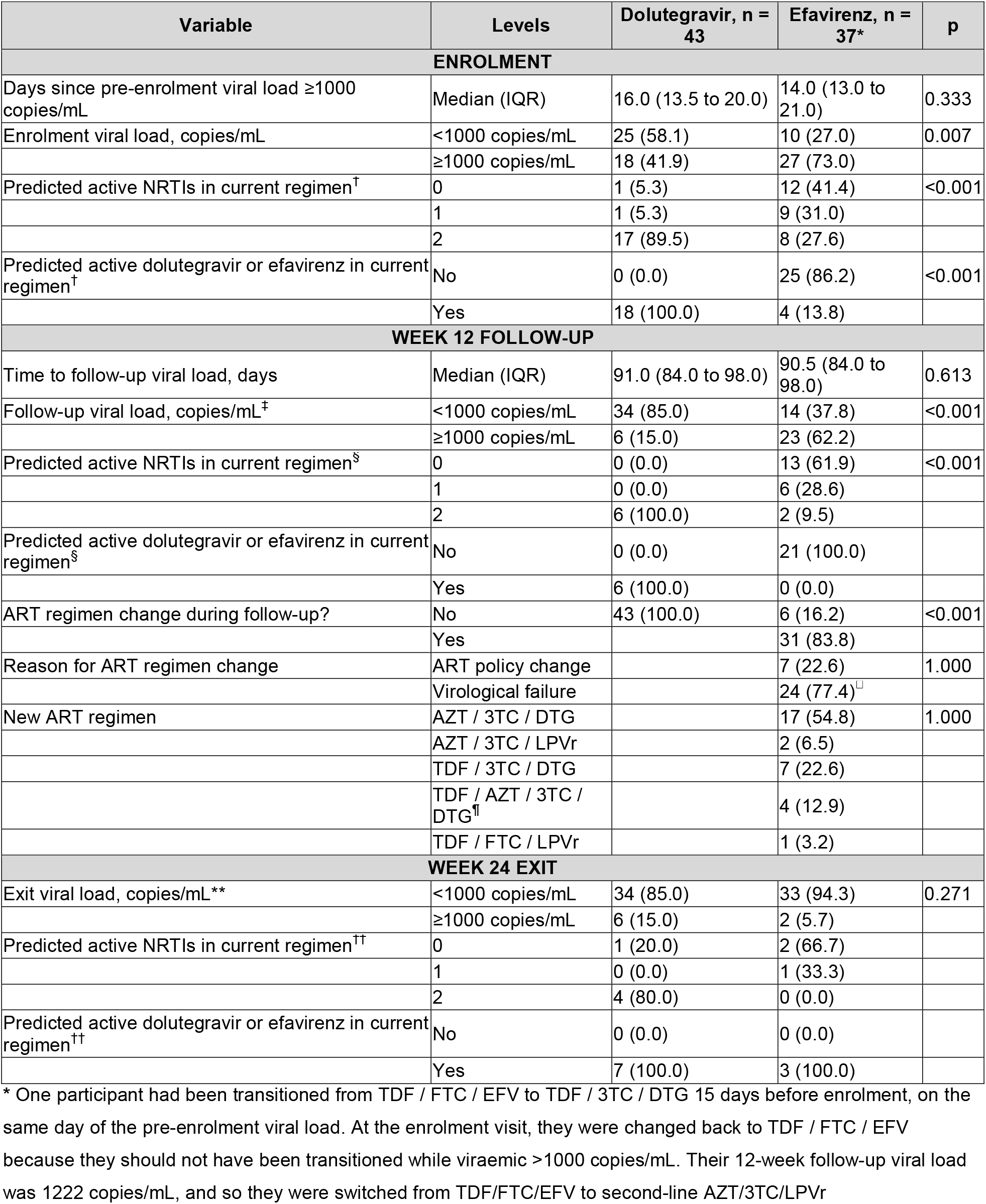
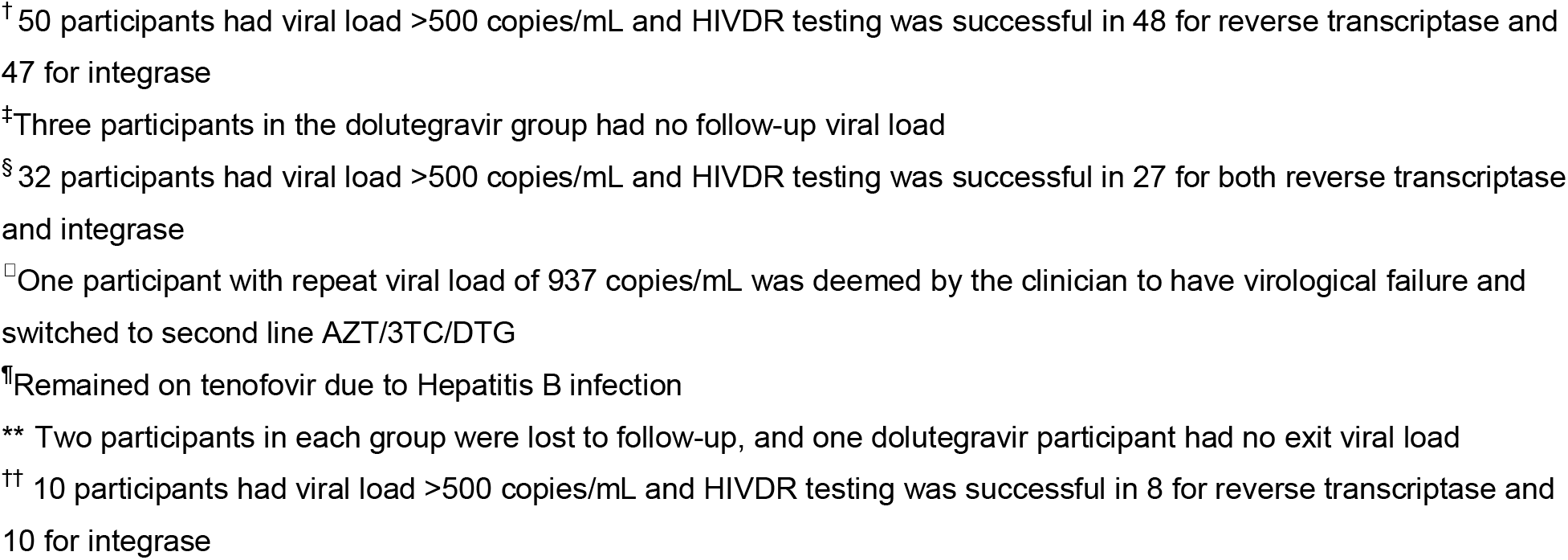
Outcomes among patients with viraemia on dolutegravir and efavirenz based first-line ART.

| Variable | Levels | Dolutegravir, n = 43 | Efavirenz, n = 37* | p |
| --- | --- | --- | --- | --- |
| ENROLMENT |  |  |  |  |
| Days since pre-enrolment viral load ≥1000 copies/mL | Median (IQR) | 16.0 (13.5 to 20.0) | 14.0 (13.0 to 21.0) | 0.333 |
| Enrolment viral load, copies/mL | <1000 copies/mL | 25 (58.1) | 10 (27.0) | 0.007 |
|  | ≥1000 copies/mL | 18 (41.9) | 27 (73.0) |  |
| Predicted active NRTIs in current regimen<sup>†</sup> | 0 | 1 (5.3) | 12 (41.4) | <0.001 |
|  | 1 | 1 (5.3) | 9 (31.0) |  |
|  | 2 | 17 (89.5) | 8 (27.6) |  |
| Predicted active dolutegravir or efavirenz in current regimen<sup>†</sup> | No | 0 (0.0) | 25 (86.2) | <0.001 |
|  | Yes | 18 (100.0) | 4 (13.8) |  |
| WEEK 12 FOLLOW-UP |  |  |  |  |
| Time to follow-up viral load, days | Median (IQR) | 91.0 (84.0 to 98.0) | 90.5 (84.0 to 98.0) | 0.613 |
| Follow-up viral load, copies/mL<sup>‡</sup> | <1000 copies/mL | 34 (85.0) | 14 (37.8) | <0.001 |
|  | ≥1000 copies/mL | 6 (15.0) | 23 (62.2) |  |
| Predicted active NRTIs in current regimen<sup>§</sup> | 0 | 0 (0.0) | 13 (61.9) | <0.001 |
|  | 1 | 0 (0.0) | 6 (28.6) |  |
|  | 2 | 6 (100.0) | 2 (9.5) |  |
| Predicted active dolutegravir or efavirenz in current regimen<sup>§</sup> | No | 0 (0.0) | 21 (100.0) |  |
|  | Yes | 6 (100.0) | 0 (0.0) |  |
| ART regimen change during follow-up? | No | 43 (100.0) | 6 (16.2) | <0.001 |
|  | Yes |  | 31 (83.8) |  |
| Reason for ART regimen change | ART policy change |  | 7 (22.6) | 1.000 |
|  | Virological failure |  | 24 (77.4)<sup>□</sup> |  |
| New ART regimen | AZT / 3TC / DTG |  | 17 (54.8) | 1.000 |
|  | AZT / 3TC / LPVr |  | 2 (6.5) |  |
|  | TDF / 3TC / DTG |  | 7 (22.6) |  |
|  | TDF / AZT / 3TC / DTG<sup>¶</sup> |  | 4 (12.9) |  |
|  | TDF / FTC / LPVr |  | 1 (3.2) |  |
| WEEK 24 EXIT |  |  |  |  |
| Exit viral load, copies/mL** | <1000 copies/mL | 34 (85.0) | 33 (94.3) | 0.271 |
|  | ≥1000 copies/mL | 6 (15.0) | 2 (5.7) |  |
| Predicted active NRTIs in current regimen<sup>††</sup> | 0 | 1 (20.0) | 2 (66.7) |  |
|  | 1 | 0 (0.0) | 1 (33.3) |  |
|  | 2 | 4 (80.0) | 0 (0.0) |  |
| Predicted active dolutegravir or efavirenz in current regimen<sup>††</sup> | No | 0 (0.0) | 0 (0.0) |  |
|  | Yes | 7 (100.0) | 3 (100.0) |  |

\* One participant had been transitioned from TDF / FTC / EFV to TDF / 3TC / DTG 15 days before enrolment, on the same day of the pre-enrolment viral load. At the enrolment visit, they were changed back to TDF / FTC / EFV because they should not have been transitioned while viraemic $>1000$ copies/mL. Their 12-week follow-up viral load was 1222 copies/mL, and so they were switched from TDF/FTC/EFV to second-line AZT/3TC/LPVr
† 50 participants had viral load >500 copies/mL and HIVDR testing was successful in 48 for reverse transcriptase and 47 for integrase
‡ Three participants in the dolutegravir group had no follow-up viral load
§ 32 participants had viral load >500 copies/mL and HIVDR testing was successful in 27 for both reverse transcriptase and integrase
□ One participant with repeat viral load of 937 copies/mL was deemed by the clinician to have virological failure and switched to second line AZT/3TC/DTG
¶ Remained on tenofovir due to Hepatitis B infection
†† 10 participants had viral load >500 copies/mL and HIVDR testing was successful in 8 for reverse transcriptase and 10 for integrase

#### Follow-up

By the time of the 12-week VL, participants in both the dolutegravir and efavirenz group had a median of 1 (IQR 1, 1) enhanced adherence counselling sessions. Only 6/43 (15.0%) of dolutegravir participants had a VL ≥1000 copies/mL and were classified as having viral failure, compared to 23/37 (62.2%) efavirenz participants (p<0.001). Of the 27/32 with VL >500 copies/mL and successful HIVDR testing, none of the dolutegravir participants had dolutegravir or NRTI DRMs, compared to 19/21 (90.5%, 69.6, 98.4) efavirenz participants who had resistance against the NRTI backbone (p<0.001). 21/21 (100%, 81.4, 100) had resistance against efavirenz. All 23 efavirenz participants with confirmed viral failure at 12 weeks, and one other with a repeat VL of 937 copies/mL, were switched to second-line regimens (Tables 1 and S2), at a median of 90 days (IQR 84, 99) weeks after enrolment. The commonest second line regimen was zidovudine, lamivudine and dolutegravir.

At the 24-week exit visit, two participants in each group were lost to follow-up, and one dolutegravir participant had no exit viral load taken. Of those with exit viral loads, viraemia was detected in 6/40 (15.0%) of those who were receiving dolutegravir at enrolment, versus 2/35 (5.7%) of those who were receiving efavirenz at enrolment (p=0.271). Among the 8/10 with successful NRTI HIVDR testing, 1/5 (20.0%, 2.5, 64.1) in the dolutegravir enrolment group had resistance against the NRTI backbone versus 3/3 (100%, 40.0, 100) in the efavirenz enrolment group. One participant who was receiving TLD from enrolment, and had only NNRTI DRMs at enrolment, developed an emergent K65R mutation by week 24 (Table S2). There were no dolutegravir DRMs detected from enrolment to study exit in any participants.

## DISCUSSION

At enrolment and 12-week follow-up, people receiving efavirenz-based ART with viraemia had high levels of DRMs against their first-line regimen, while people receiving dolutegravir had minimal resistance. Consequently dolutegravir participants had higher levels of resuppression at 12-weeks compared to efavirenz. After switching to second-line ART, 24-week viral resuppression in efavirenz participants became similar to dolutegravir, with few DRMs in both groups.

Among participants receiving dolutegravir-based ART at baseline, there were no INSTI mutations, meaning that viraemia was likely caused by poor adherence. Our study is one of the first to report outcomes among people experiencing viraemia on first-line dolutegravir in LMICs, with 85.0% achieving viral suppression <1000 copies/mL after 12 weeks. In contrast, a high proportion of participants receiving efavirenz-based ART had baseline NRTI and NNRTI resistance, meaning resistance was contributing to viraemia. After 12 weeks, 37.8% re-suppressed to <1000 copies/mL, similar to the 46.4% among people receiving NNRTI-based ART in a large systematic review.^11^ The remaining participants only resuppressed after switching to second-line ART. One other study compares resuppression among people with viraemia receiving dolutegravir versus efavirenz in LMICs.^12^ Among people with viraemia after initiating ART in the ADVANCE trial, resuppression was more frequent in the dolutegravir group (155/247, 62.8%) compared to efavirenz (44/138, 32%, p <0.001). There was one case of emergent resistance to dolutegravir.

Strengths of our study include the focus on people with viraemia while receiving dolutegravir, successful HIVDR testing in a high proportion of those with viraemia, and frequent VL testing. The small sample size meant we could not adjust for potential confounding factors that could contribute the difference in outcomes between dolutegravir and efavirenz participants. For example, people who were transitioned to dolutegravir may be better engaged in care or motivated to adhere to treatment, and therefore also more likely to resuppress. Follow-up time was short, and the median time on dolutegravir was less than a year.

Nevertheless, our findings, alongside those of the ADVANCE study, demonstrate that early in the South African rollout, viraemia among people receiving dolutegravir is largely due to poor adherence, rather than drug resistance. This supports the current South African and WHO guidelines, which do not recommend early switching to second-line ART among people receiving dolutegravir with viraemia. The high cost of HIV drug resistance testing, and low prevalence of drug resistance, mean that South African Guidelines only recommend drug resistance after two years of viraemia. Further evidence is needed to determine the extent and impact of emergent DRMs with longer term viraemia on dolutegravir. In the meantime, managing viraemia among people receiving dolutegravir should have a renewed focus on interventions to support adherence, rather than managing HIVDR.

## Supporting information

see supplementary material

## Data Availability

Bona fide researchers will be able to request access to anonymised trial data by contacting the corresponding author.

## LIST OF ABBREVIATIONS

ART: antiretroviral therapy
PLHIV: people living with HIV
LMIC: low- and middle-income countries
UNAIDS: Joint United Nations Programme on HIV/AIDS
WHO: World Health Organization

## Author contributions

JD and NG conceived the study. YS, RL, EB, PM, NS, PKD, GH and CB contributed to study design and implementation. JD analysed the data and wrote the first draft of the manuscript. All authors critically reviewed and edited the manuscript and consented to final publication.

## Acknowledgements

The authors would like to thank all participants in the study and acknowledge the work and support of staff at the Prince Cyril Zulu Clinic, Mafakathini Clinic, eThekwini Municipality, CAPRISA and the National Health Laboratory Services at Addington and Inkosi Albert Luthuli Hospitals.

## REFERENCES

1. Dorward J, Lessells R, Drain PK, et al. Dolutegravir for first-line antiretroviral therapy in lowincome and middle-income countries: uncertainties and opportunities for implementation and research. Lancet HIV 2018;5(7):e400–e04. doi: 10.1016/S2352-3018(18)30093-6 [published Online First: 2018/06/10]

2. Vitoria M, Hill A, Ford N, et al. The transition to dolutegravir and other new antiretrovirals in low-income and middle-income countries: what are the issues? AIDS 2018;32(12):1551–61. doi: 10.1097/QAD.0000000000001845 [published Online First: 2018/05/11]

3. Kanters S, Vitoria M, Zoratti M, et al. Comparative efficacy, tolerability and safety of dolutegravir and efavirenz 400mg among antiretroviral therapies for first-line HIV treatment: A systematic literature review and network meta-analysis. EClinicalMedicine 2020;28:100573. doi: 10.1016/j.eclinm.2020.100573 [published Online First: 2020/12/10]

4. Hamers RL, Rinke de Wit TF, Holmes CB. HIV drug resistance in low-income and middleincome countries. The Lancet HIV 2018;5(10):e588–e96. doi: 10.1016/S2352-3018(18)30173-5

5. Gregson J, Tang M, Ndembi N, et al. Global epidemiology of drug resistance after failure of WHO recommended first-line regimens for adult HIV-1 infection: a multicentre retrospective cohort study. The Lancet Infectious Diseases 2016;16(5):565–75. doi: 10.1016/S1473-3099(15)00536-8

6. Hunt GM, Dokubo EK, Takuva S, et al. Rates of virological suppression and drug resistance in adult HIV-1-positive patients attending primary healthcare facilities in South Africa. Journal of Antimicrobial Chemotherapy 2017;72:3141–48. doi: 10.1093/jac/dkx252

7. Dorward J, Sookrajh Y, Ngobese H, et al. Protocol for a randomised feasibility study of Point-Of-care HIV viral load testing to Enhance Re-suppression in South Africa: the POwER study. BMJ Open 2021;11(2):e045373. doi: 10.1136/bmjopen-2020-045373

8. Dorward J, Sookrajh Y, Lessells RJ, et al. Point-of-care viral load testing to manage HIV viraemia during the rollout of dolutegravir-based ART in South Africa: a randomised feasibility study (POwER) Journal of Acquired Immune Deficiency Syndromes 2023

9. The South African National Department of H. 2019 ART Clinical Guidelines for the management of HIV in Adults, Pregnancy, Adolescents, Children, Infants and Neonates. Pretoria, South Africa, 2019.

10. South African National Department of Health. Adherence Guidelines for HIV, TB and NCDs: Updated March 2020 Pretoria, South Africa 2020 [Available from: https://www.knowledgehub.org.za/elibrary/adherence-guidelines-hiv-tb-and-ncds-standard-operating-procedures-2020.

11. Ford N, Orrell C, Shubber Z, et al. HIV viral resuppression following an elevated viral load: a systematic review and meta-analysis. Journal of the International AIDS Society 2019;22(11):1–6. doi: 10.1002/jia2.25415

12. Pepperrell T, Venter WDF, McCann K, et al. Participants on Dolutegravir Resuppress Human Immunodeficiency Virus RNA After Virologic Failure: Updated Data from the ADVANCE Trial. Clin Infect Dis 2021;73(4):e1008–e10. doi: 10.1093/cid/ciab086

