## Supplementary material for "Viraemia and HIV drug resistance among people receiving dolutegravir versus efavirenz-based first-line antiretroviral therapy": see supplementary material

**SUPPLEMENTARY FILE**

### 1. Details of HIV drug resistance testing

We attempted sequencing of HIV-1 *pol* (protease [PR], reverse transcriptase [RT] and integrase [IN]) for all samples with VL ≥500 copies/mL. Following RNA extraction, we amplified PR, RT and IN genes using the amplification module of the Applied Biosystems HIV-1 Genotyping Kit with Integrase (Thermo Fisher Scientific, Waltham, MA); and sequenced on the Illumina MiSeq platform (Illumina, San Diego, CA). We identified drug resistance mutations (DRMs) at >20% frequency using Stanford HIVdb (version 9.1).

### 2. Table S1: Baseline characteristics by ART regimen at enrolment

| **Variable** | **Levels** | **Dolutegravir,**  **n = 43** | **Efavirenz,**  **n = 37** | **Total** |
| --- | --- | --- | --- | --- |
| Age, years | Median (IQR) | 40.0 (34.0 to 48.0) | 38.0 (33.0 to 41.0) | 38.5 (33.0 to 45.0) |
| Gender | Female | 24 (55.8) | 23 (62.2) | 47 (58.8) |
|  | Male | 19 (44.2) | 14 (37.8) | 33 (41.2) |
| Time since ART initiation, years | Median (IQR) | 2.5 (0.6 to 6.0) | 4.1 (1.4 to 5.2) | 3.2 (1.0 to 6.0) |
| NRTI backbone | ABC | 1 (2.3) |  | 1 (1.2) |
|  | TDF | 42 (97.7) | 37 (100.0) | 79 (98.8) |
| Time on current regimen, years | Median (IQR) | 0.7 (0.5 to 1.1) | 3.2 (1.1 to 5.0) | 1.1 (0.6 to 2.9) |
| History of other chronic medical conditions | No | 37 (86.0) | 34 (91.9) | 71 (88.8) |
|  | Yes | 6 (14.0) | 3 (8.1) | 9 (11.2) |
| Current ART side effects | No | 41 (95.3) | 37 (100.0) | 78 (97.5) |
|  | Yes | 2 (4.7) |  | 2 (2.5) |
| Last time participant missed a dose of ART | Within past the week | 10 (23.3) | 7 (18.9) | 17 (21.2) |
|  | 1-2 weeks ago | 3 (7.0) | 1 (2.7) | 4 (5.0) |
|  | 2-4 weeks ago | 7 (16.3) | 5 (13.5) | 12 (15.0) |
|  | 1-3 months ago | 5 (11.6) | 5 (13.5) | 10 (12.5) |
|  | >3 months ago | 1 (2.3) | 1 (2.7) | 2 (2.5) |
|  | Never | 17 (39.5) | 18 (48.6) | 35 (43.8) |
| Hazardous drinking (AUDIT-C) | No | 30 (69.8) | 24 (64.9) | 54 (67.5) |
|  | Yes | 13 (30.2) | 13 (35.1) | 26 (32.5) |
| Smoking status | Current smoker | 8 (18.6) | 5 (13.5) | 13 (16.2) |
|  | Never smoked | 35 (81.4) | 31 (83.8) | 66 (82.5) |
|  | Ex-smoker |  | 1 (2.7) | 1 (1.2) |
| Body mass index, kg/m2 | Median (IQR) | 28.4 (24.5 to 32.2) | 25.4 (22.2 to 31.5) | 27.3 (22.9 to 31.7) |
| Ethnicity | Black African | 43 (100.0) | 36 (97.3) | 79 (98.8) |
|  | Other |  | 1 (2.7) | 1 (1.2) |
| Highest level of education completed | None | 0 (0.0) | 0 (0.0) | 0 (0.0) |
|  | Primary school only | 1 (2.3) | 0 (0.0) | 1 (1.2) |
|  | Secondary school but not matric | 22 (51.2) | 17 (45.9) | 39 (48.8) |
|  | Matriculation | 16 (37.2) | 16 (43.2) | 32 (40.0) |
|  | Tertiary | 4 (9.3) | 4 (10.8) | 8 (10.0) |
| Employment status | Unemployed | 13 (30.2) | 15 (40.5) | 28 (35.0) |
|  | Informal employment | 5 (11.6) | 1 (2.7) | 6 (7.5) |
|  | Part time employment | 7 (16.3) | 2 (5.4) | 9 (11.2) |
|  | Full-time employment | 15 (34.9) | 12 (32.4) | 27 (33.8) |
|  | Student | 1 (2.3) | 4 (10.8) | 5 (6.2) |
|  | Self-employed | 2 (4.7) | 3 (8.1) | 5 (6.2) |
| Monthly personal income, ZAR | <1000 | 16 (37.2) | 20 (54.1) | 36 (45.0) |
|  | 1000-4000 | 14 (32.6) | 9 (24.3) | 23 (28.8) |
|  | 4001-8000 | 8 (18.6) | 8 (21.6) | 16 (20.0) |
|  | >8000 | 5 (11.6) | 0 (0.0) | 5 (6.2) |
| Enrolment CD4 count category, cells/µL | <200 | 6 (14.0) | 15 (40.5) | 21 (26.2) |
|  | 200-349 | 12 (27.9) | 9 (24.3) | 21 (26.2) |
|  | 350-499 | 14 (32.6) | 4 (10.8) | 18 (22.5) |
|  | >=500 | 11 (25.6) | 9 (24.3) | 20 (25.0) |

### 3. Table S2: HIV drug resistance mutations among participants with viraemia*

| **ENROLMENT** | | | | **FOLLOW-UP** | | | | **EXIT** | | |
| --- | --- | --- | --- | --- | --- | --- | --- | --- | --- | --- |
| **ART** | **VL** | **NRTI mutations** | **NNRTI mutations** | **VL** | **NRTI mutations** | **NNRTI mutations** | **New Regimen** | **VL** | **NRTI mutations** | **NNRTI mutations** |
| TEE | 3530 | None | V108I | 91 | NA | NA | TLD | 100 | NA | NA |
| TEE | 2550 | None | None | 231 | NA | NA | Not switched | <20 | NA | NA |
| TEE | 128000 | A62V,K65R,M184V | L100I,K103N,Y188L | 5490000 | A62V,K65R,M184V | L100I,K103N,V106VI,Y188L | ZLD | 384 | NA | NA |
| TEE | 1450 | K219E | None | 82600 | K65KR,M184MIV | K103N,V106M | ZLD | <20 | NA | NA |
| TEE | 13500 | D67N,K70E,M184V | K101E,V106M,G190A,F227L | 69276 | D67N,K70EQ,M184V | K101E,V106M,G190A,F227L | ZLD | 64.7 | NA | NA |
| TEE | 116000 | D67N,M184V,K219R | A98G,K103N,P225H | 124215 | K65R,M184I | K103N,L234I | TDF / ZLD | 41.7 | NA | NA |
| TEE | 6320 | K65R,D67G | V106M,G190A | 9020 | A62V,K65R,D67G,Y115F,M184V | V106M,G190A | ZLD | <20 | NA | NA |
| TEE | 10900 | M184V | K101P,K103N | 5000 | M184V | K101P,K103N | ZLD | TND | NA | NA |
| TEE^†^ | 939 | K65R,M184V | A98G,K101E,V108I,Y181C | 1222 | HIVDR testing unsuccessful | HIVDR testing unsuccessful | AZT / 3TC / LPVr | <20 | NA | NA |
| TEE | 6410 | K70R,M184V,K219Q | K103N,P225H,K238T | 2030 | HIVDR testing unsuccessful | HIVDR testing unsuccessful | AZT / 3TC / LPVr | 85 | NA | NA |
| TEE | 8810 | D67N,K70E,L74I,M184V | K103N,V106M,V179T | 3638 | D67N,K70E,L74LI,M184V | K103N,V106M,V179T | ZLD | <20 | NA | NA |
| TEE | 59500 | K65R,D67N,M184I | A98G,K103N,L234I | 20839 | None | K103N | ZLD | 99.5 | NA | NA |
| TEE | 94100 | L74I,M184V | K103N,G190S | 21600 | L74LI,M184V | K103N,V179IT,G190S | TDF / ZLD | NA | NA | NA |
| TEE | 284000 | K70E,L74I,M184V,K219R | A98G,K103N,P225H,F227L | 419846 | K70E,M184V,K219R | A98G,K103N,P225H,F227L | TDF / ZLD | 42.4 | NA | NA |
| TEE | 1190 | K219R | K103N | 0 | NA | NA | TLD | TND | NA | NA |
| TEE | 5130 | A62V,K65R,K70Q,M184V | K101P,Y181C,G190A | 1388 | A62V,K65R,K70Q,M184V | K101P,Y181C,G190A | ZLD | <20 | NA | NA |
| TEE | 439 | NA | NA | 937 | A62AV,K65R,K70T,M184V | V106M,V108I,Y188C,F227L | ZLD | TND | NA | NA |
| TEE | 297 | NA | NA | 1390 | K65R,M184V | L100I,K103N | ZLD | TND | NA | NA |
| TEE | 352000 | T69D,M184V | V179D,Y188L | 235000 | T69D,M184V | V179D,Y188L | ZLD | 23.7 | NA | NA |
| TEE | 1420 | D67N,M184V,K219R | K103N,V108I | 615 | HIVDR testing unsuccessful | HIVDR testing unsuccessful | Not switched | NA | NA | NA |
| TEE | 6320 | D67T,K70R,M184V,T215V,K219E | A98G,K103N,V108I,P225H | 10934 | D67T,K70R,M184V,T215V,K219E | A98G,K103N,V108I,P225H | TDF / ZLD | TND | NA | NA |
| TEE | 22300 | K65R | V106M,V179D | 1630 | M184V | V106M,V179D | ZLD | <20 | NA | NA |
| TEE | 534 | HIVDR testing unsuccessful | HIVDR testing unsuccessful | 4690 | HIVDR testing unsuccessful | HIVDR testing unsuccessful | TDF / FTC / LPVr | TND | NA | NA |
| TEE | 18900 | A62V,K65R,K70T,M184V | V106M,V179D,Y181C,H221Y,F227I | 64000 | A62V,K65R,K70T,M184V | V106M,V179D,Y181C,H221Y,F227I | ZLD | TND | NA | NA |
| TEE | 1180 | None | K103N,V106M,G190A | 7140 | M184V | K103N,V106M,G190A | ZLD | <20 | NA | NA |
| TEE | 2200 | D67N,M184V,K219E | K103N,V108I | 2225 | D67N,K70R,M184V,K219E | K103N,V108I | ZLD | 109 | NA | NA |
| TEE | 267000 | None | V106M | 586 | None | V106M | TLD | TND | NA | NA |
| TEE | 21900 | None | None | 40 | NA | NA | TLD | TND | NA | NA |
| TEE | 1360 | K70R,M184V,K219Q | K103N,P225H,K238T | 251 | NA | NA | TLD | TND | NA | NA |
| TEE | 6050 | K70Q,L74I,M184V | K103S,V106M | 422320 | K70KQ,L74I,M184V | K103NS,V106M | ZLD | 5810 | K70KQ,L74LI,M184V | K103NS,V106M |
| TEE | 20100 | None | K103N | 1660 | HIVDR testing unsuccessful | HIVDR testing unsuccessful | ZLD | 4370 | D67N,M184V,K219E | K103N,P225H |
| TEE | 959 | M41L,M184V,T215Y | A98AG,K103N,E138Q,P225H | 191 | NA | NA | TLD | 802 | M41L,M184V,T215Y | K103N,E138Q,P225H |
| TLD | 177827 | NA | NA | 148000 | None | None | Not switched | NA | NA | NA |
| TLD | 177827 | NA | NA | 148000 | None | None | Not switched | NA | NA | NA |
| TLD | 18600 | None | K103N | 84 | NA | NA | Not switched | 22.1 | NA | NA |
| TLD | 2180 | None | None | NA | NA | NA | Not switched | <20 | NA | NA |
| TLD | 515 | None | V106M,V179VD | 0 | NA | NA | Not switched | 23.9 | NA | NA |
| TLD | 3950 | M184V | K103N,V106M | 40 | NA | NA | Not switched | 82.9 | NA | NA |
| TLD | 27 | NA | NA | 41 | NA | NA | Not switched | 7980 | None | None |
| TLD | 738 | None | None | 40 | NA | NA | Not switched | 24.3 | NA | NA |
| TLD | 13400 | None | None | 0 | NA | NA | Not switched | 220 | NA | NA |
| TLD | 327 | NA | NA | 20 | NA | NA | Not switched | 676 | HIVDR testing unsuccessful | HIVDR testing unsuccessful |
| TLD | 18000 | None | None | 86700 | None | None | Not switched | TND | NA | NA |
| TLD | 4910 | None | None | 20 | NA | NA | Not switched | TND | NA | NA |
| TLD | 2980 | None | Y188L | 40 | NA | NA | Not switched | 52600 | None | Y188L |
| TLD | 333000 | None | None | 34 | NA | NA | Not switched | TND | NA | NA |
| TLD | 296000 | None | None | 40 | NA | NA | Not switched | 33.3 | NA | NA |
| TLD | <20 | NA | NA | 1280 | None | None | Not switched | TND | NA | NA |
| TLD | 11700 | None | None | 40 | NA | NA | Not switched | TND | NA | NA |
| TLD | 14900 | None | K103N,E138A | 72 | NA | NA | Not switched | 16900 | K65KR | K103N,E138A |
| TLD | 3260 | None | Y188L | 5760 | None | Y188L | Not switched | 47600 | None | Y188L |
| TLD | 238 | NA | NA | 40 | NA | NA | Not switched | 10100 | None | None |
| TLD | 23400 | None | None | 50 | NA | NA | Not switched | NA | NA | NA |
| TLD | 215000 | None | A98G | 25 | NA | NA | Not switched | 33.9 | NA | NA |
| TLD | 35800 | None | A98G | 1215 | None | None | Not switched | 25.10 | NA | NA |
| TLD | 48.1 | NA | NA | NA | NA | NA | Not switched | 3520 | HIVDR testing unsuccessful | HIVDR testing unsuccessful |
| TLD | 3600 | None | V106M,Y188C | 53 | NA | NA | Not switched | 42.2 | NA | NA |
| ALD | 95100 | L74I,M184V | K103N,G190S | 6243 | None | None | Not switched | <20 | NA | NA |

* No major PI or INSTI mutations detected throughout follow-up. ^†^ One participant had been transitioned from TDF / FTC / EFV to TDF / 3TC / DTG 15 days before enrolment, on the same day that the pre-enrolment viral load was viraemic. At the enrolment visit, they were changed back to TDF / FTC / EFV because they should not have been transitioned with a viral load >1000 copies/mL. TLD = tenofovir disoproxil fumarate, lamivudine and dolutegravir, ZLD = zidovudine, lamivudine and dolutegravir, TDF = tenofovir disoproxil fumarate, ALD = abacavir, lamivudine and dolutegravir. TND = not detected.
